# The effect of protein levels of ABO on pregnancy related outcomes: a Mendelian randomization study

**DOI:** 10.1101/2023.10.11.23296777

**Authors:** Yuqi Sun, Haonan Zheng, Manqing Wang, Rongrong Gu, Xueyan Wu, Qian Yang, Huiling Zhao, Yufang Bi, Jie Zheng

**Affiliations:** Department of Endocrine and Metabolic Diseases, Shanghai Institute of Endocrine and Metabolic Diseases, Ruijin Hospital, Shanghai Jiao Tong University School of Medicine, Shanghai, China; College of Health Science and Technology, Shanghai Jiao Tong University School of Medicine; Shanghai National Clinical Research Center for metabolic Diseases, Key Laboratory for Endocrine and Metabolic Diseases of the National Health Commission of the PR China, Shanghai Key Laboratory for Endocrine Tumor, Shanghai Digital Medicine Innovation Center, Ruijin Hospital, Shanghai Jiao Tong University School of Medicine, Shanghai, China; College of Basic Medical Science, Shanghai Jiao Tong University School of Medicine; MRC Integrative Epidemiology Unit (IEU), Bristol Medical School, University of Bristol, Oakfield House, Oakfield Grove, Bristol, BS8 2BN, United Kingdom

**Keywords:** Mendelian randomization, ABO, venous complication in pregnancy, gestational hypertension

## Abstract

Protein level of Histo-Blood Group ABO System Transferase (ABO) has been reported to be associated with cardiometabolic diseases. But its effect on pregnancy related outcomes was still unclear. Here we conducted a two-sample Mendelian randomization study to ascertain the putative causal roles of protein levels of ABO in pregnancy related outcomes. Cis-acting protein quantitative trait loci (pQTLs) robustly associated with protein level of ABO (P < 5x10-8) were used as instruments to proxy the ABO level (N = 35,559, data from deCODE), with two additional pQTL datasets from Fenland (N = 10,708) and INTERVAL (N = 3,301) used as validation exposures. Ten pregnancy related diseases and complications were selected as outcomes. We observed that a higher protein level of ABO showed a putative causal effect on venous complications and haemorrhoids in pregnancy (OR = 1.207, 95%CI = 1.107-1.316, colocalization probability = 91.3%), which was validated by using pQTLs from Fenland and INTERVAL. The Mendelian randomization results further showed effects of the ABO protein on gestational hypertension (OR = 0.974, 95%CI = 0.953-0.995), despite insignificance after multiple testing correction and little colocalization evidence. Sensitivity analyses, including proteome-wide Mendelian randomization of the cis-acting ABO pQTLs, showed little evidence of horizontal pleiotropy. Correctively, our study prioritised ABO as a putative causal protein for venous complications and haemorrhoids in pregnancy. Future epidemiology and clinical studies are needed to investigate whether ABO can be considered as a drug target to prevent adverse pregnancy outcomes.

## Introduction

Adverse pregnancy outcomes, e.g. pregnancy induced venous complications (PIVC), gestational hypertension, gestational oedema and preterm labor, are health conditions related to both pregnant women and newborns, and always arose concerns in our society. Recent studies indicated that preterm labor is a leading cause of perinatal morbidity and mortality in developed countries[1]. PIVC includes pregnancy related deep vein thrombosis and lower limb varicose veins, causes lower limb swelling, pain, skin ulcers, and difficulties for the daily activities (e.g. sleep), and even threatens lives of pregnant women[2]. Hemorrhoids can be considered as a common venous disease that has a significant impact on quality of life for pregnant women[3]. Gestational hypertension is a major cause of maternal, fetal and newborn morbidity and mortality, and women with pregnancy-induced hypertension syndrome are at higher risks of abruptio placentae, cerebrovascular events, organ failure and disseminated intravascular coagulation[4]. Identification of genetic regulators that influence pregnancy related outcomes will help better understand the etiology of these conditions, and may support the development of drug targets to reduce risks of these outcomes.

ABO blood group system classifies human blood into four main types: A, B, AB, and O, which indicates the presence or absence of specific antigens on the surface of red blood cells. It is genetically determined by the ABO gene (chromosome 9, q34.2), and the gene mutations in the region influence the blood group. ABO blood group is proved to be related to a wide range of diseases, especially cardiovascular ones[5]. Julia Hoglund et al. characterized the human ABO genotypes and their associations with common inflammatory and cardiovascular diseases. They found that ABO gene mutations was significantly associated with individual thrombotic risk[6]. Several studies observed the associations of ABO blood group system with pregnancy related outcomes, including stillbirth[7] and obstetrical hemorrhage risk[8]. Additionally, pregnant women with type O blood had a lower risk of cardiovascular disease comparing to those with type A, type B and type AB[9]. Yet the evidence supporting the association between ABO blood group and pregnancy related outcomes is still limited, requiring large birth cohorts or record-linkage data. This inspired us to further investigate the function of plasma protein level of ABO (i.e., Histo-Blood Group ABO System Transferase) on top of the traditional blood tests, in order to understand the relationships between the ABO level and pregnancy related outcomes. Given it would not be feasible to conduct a randomized control trail among pregnant women due to ethical concerns[10], Mendelian randomization (MR) provides an alternative way to assess the impact of ABO protein level on pregnancy outcomes. Two-sample MR is a genetic epidemiology method using germline genetic variants (also called single nucleotide polymorphisms (SNPs)) as instruments to estimate the causal effect of an exposure on an outcome[11]. Previous MR studies identified effects of protein level of ABO on many diseases, including COVID-19[12] and ischemic stroke[13]. However, its casual relationships with pregnancy related outcomes remain unclear. Recently available largescale genome-wide association studies (GWASs) of protein level of ABO[14-16], and adverse pregnancy outcomes[17-19] provided a timely opportunity to investigate the causal effect of the ABO level on those outcomes using MR.

The aim of this study was to estimate the casual associations of protein level of ABO with ten pregnancy related outcomes using two-sample MR. To increase reliability of the findings, the causal estimates were validated in three independent ABO datasets (deCODE[14], Fenland[15] and INTERVAL[16]). To further increase the possibility of identifying true causality, we applied a sensitivity analysis pipeline[20], including Steiger filtering[21], and genetic colocalization[22-24]. Given pleiotropic potential of the ABO instruments, we applied a proteome-wide MR to further investigate the possibility of horizontal pleiotropy.

## Results

In this study, MR analyses and Steiger filtering tests were performed using the TwoSampleMR package, and colocalization analysis were conducted using the coloc package. Results were visualized using the ggplot2 and LocusZoom[25] in the R software platform (version 4.2.2; R Development Core Team).

### Summary of ABO instrument selection

We selected two instruments (rs1053878 and rs950529388) associated with protein level of ABO that reached the genome-wide significance (P-value < 5x10−8) in the discovery cohort (i.e., deCODE). The instrument strength of a combination of these two instruments were strong (total F statistic = 18667.266), where each of the individual instrument also showed a strong strength (Table S1A). The two ABO instruments were weakly correlated with each other (LD r2 < 0.3), and were used in the discovery MR analyses (Table S4A). Other two sets of instruments (two instruments selected from Fenland and one instruments selected from INTERVAL) were used to proxy ABO protein level in the validation MR analyses (Table S2 and S3). All the five selected instruments were common variants with a minor allele frequency > 0.05, passed the Steiger filtering test of directionality (Table S7A, S7B and S7C) and were not palindromic SNPs with minor allele frequencies larger than 0.42.

### Effect of protein levels of ABO on pregnancy related outcomes using deCODE data

Figure 2 visualizes the pregnancy outcomes achieving statistical significance before FDR correction. Table 1 and Figure 3 present two-sample MR estimates for associations of ABO pQTL in deCODE with pregnancy-related outcomes. In the discovery analysis, MR suggested that higher protein levels of ABO showed a putative causal effect on increased risks of pregnancy induced venous complications and haemorrhoids in pregnancy (OR = 1.207; 95% confidence interval [95% CI]: 1.107–1.316; FDR-adjusted P-value = 2.008 x 10-4) (Figure 3). Strong evidence of colocalization indicated that venous complications and haemorrhoids in pregnancy (colocalization probability = 91.3%) shared the same causal variants with the protein level of ABO (Table 1). The MR analyses for other pregnancy related outcomes did not reach the FDR threshold, and their colocalization analyses further suggested little evidence of causal effects on those outcomes. Through the LD check between ABO pQTLs and venous complications, we identified that one SNP (rs687621, merged with our instrument -rs950529388) in deCODE was strongly correlated with the leading SNP in the outcome dataset (LD r2 > 0.8), suggesting a potential causal relationship. However, LD check for the other instrument (rs1053878) did not show such a relationship (Figure 4).

**Table 1.**
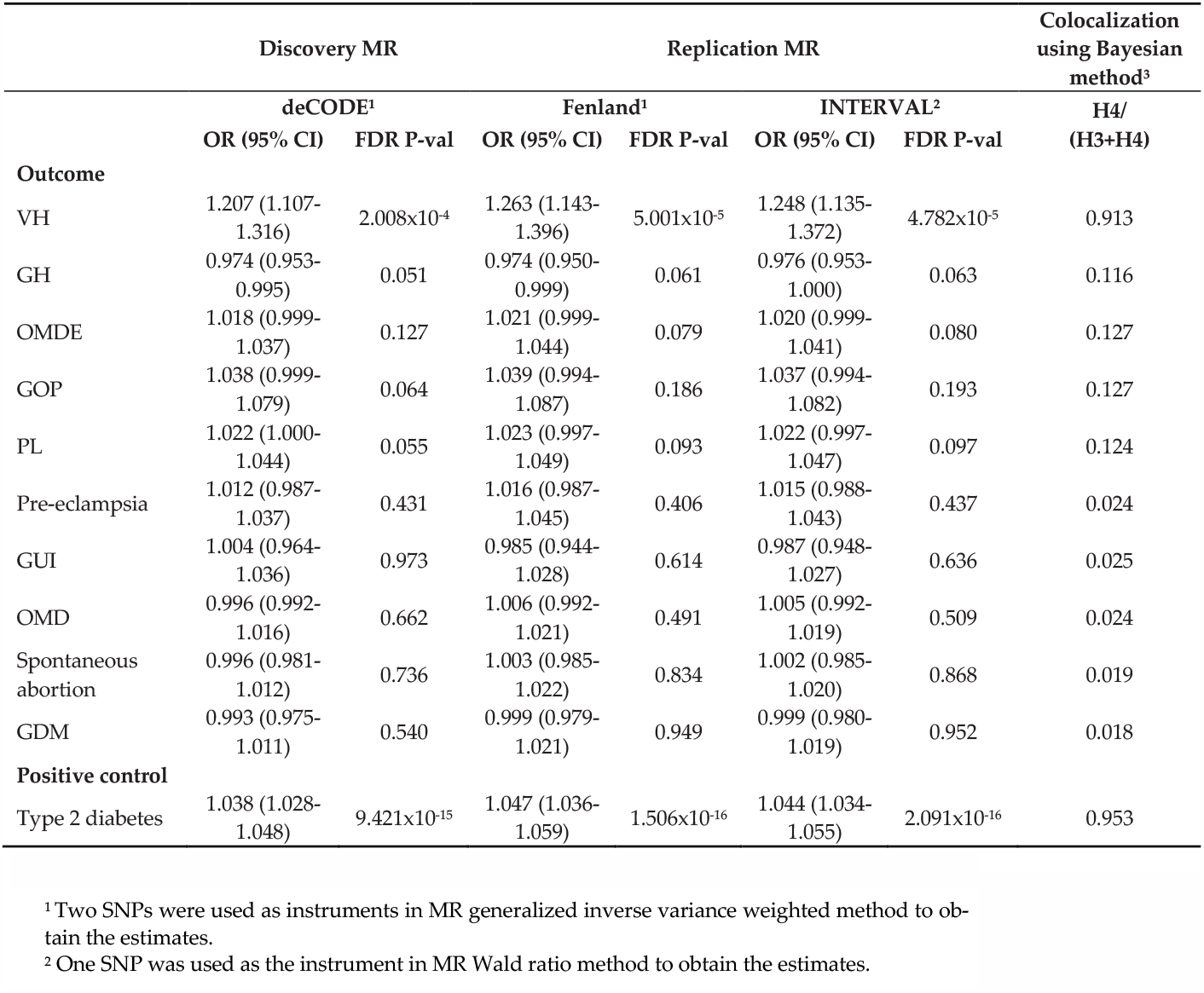
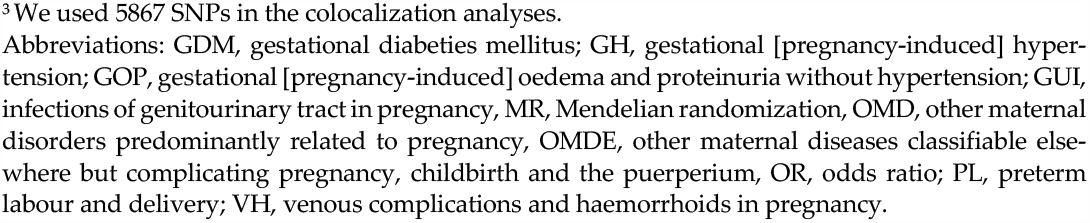
Two-sample MR estimates for associations of ABO protein with ten pregnancy outcomes using instruments from deCODE, Fenland and INTERVAL.

| Outcome | Discovery MR |  | Replication MR |  |  |  | Colocalization using Bayesian method<sup>3</sup> |
| --- | --- | --- | --- | --- | --- | --- | --- |
|  | deCODE<sup>1</sup> |  | Fenland<sup>1</sup> |  | INTERVAL<sup>2</sup> |  | H4/ (H3+H4) |
|  | OR (95% CI) | FDR P-val | OR (95% CI) | FDR P-val | OR (95% CI) | FDR P-val |  |
| VH | 1.207 (1.107-1.316) | 2.008x10<sup>-4</sup> | 1.263 (1.143-1.396) | 5.001x10<sup>-5</sup> | 1.248 (1.135-1.372) | 4.782x10<sup>-5</sup> | 0.913 |
| GH | 0.974 (0.953-0.995) | 0.051 | 0.974 (0.950-0.999) | 0.061 | 0.976 (0.953-1.000) | 0.063 | 0.116 |
| OMDE | 1.018 (0.999-1.037) | 0.127 | 1.021 (0.999-1.044) | 0.079 | 1.020 (0.999-1.041) | 0.080 | 0.127 |
| GOP | 1.038 (0.999-1.079) | 0.064 | 1.039 (0.994-1.087) | 0.186 | 1.037 (0.994-1.082) | 0.193 | 0.127 |
| PL | 1.022 (1.000-1.044) | 0.055 | 1.023 (0.997-1.049) | 0.093 | 1.022 (0.997-1.047) | 0.097 | 0.124 |
| Pre-eclampsia | 1.012 (0.987-1.037) | 0.431 | 1.016 (0.987-1.045) | 0.406 | 1.015 (0.988-1.043) | 0.437 | 0.024 |
| GUI | 1.004 (0.964-1.036) | 0.973 | 0.985 (0.944-1.028) | 0.614 | 0.987 (0.948-1.027) | 0.636 | 0.025 |
| OMD | 0.996 (0.992-1.016) | 0.662 | 1.006 (0.992-1.021) | 0.491 | 1.005 (0.992-1.019) | 0.509 | 0.024 |
| Spontaneous abortion | 0.996 (0.981-1.012) | 0.736 | 1.003 (0.985-1.022) | 0.834 | 1.002 (0.985-1.020) | 0.868 | 0.019 |
| GDM | 0.993 (0.975-1.011) | 0.540 | 0.999 (0.979-1.021) | 0.949 | 0.999 (0.980-1.019) | 0.952 | 0.018 |
| **Positive control** |  |  |  |  |  |  |  |
| Type 2 diabetes | 1.038 (1.028-1.048) | 9.421x10<sup>-15</sup> | 1.047 (1.036-1.059) | 1.506x10<sup>-16</sup> | 1.044 (1.034-1.055) | 2.091x10<sup>-16</sup> | 0.953 |

<sup>1</sup> Two SNPs were used as instruments in MR generalized inverse variance weighted method to obtain the estimates.
<sup>2</sup> One SNP was used as the instrument in MR Wald ratio method to obtain the estimates.
<sup>3</sup> We used 5867 SNPs in the colocalization analyses.
Abbreviations: GDM, gestational diabetes mellitus; GH, gestational [pregnancy-induced] hyper- tension; GOP, gestational [pregnancy-induced] oedema and proteinuria without hypertension; GUI, infections of genitourinary tract in pregnancy, MR, Mendelian randomization, OMD, other maternal disorders predominantly related to pregnancy, OMDE, other maternal diseases classifiable else- where but complicating pregnancy, childbirth and the puerperium, OR, odds ratio; PL, preterm labour and delivery; VH, venous complications and haemorrhoids in pregnancy.

**Figure 1.**
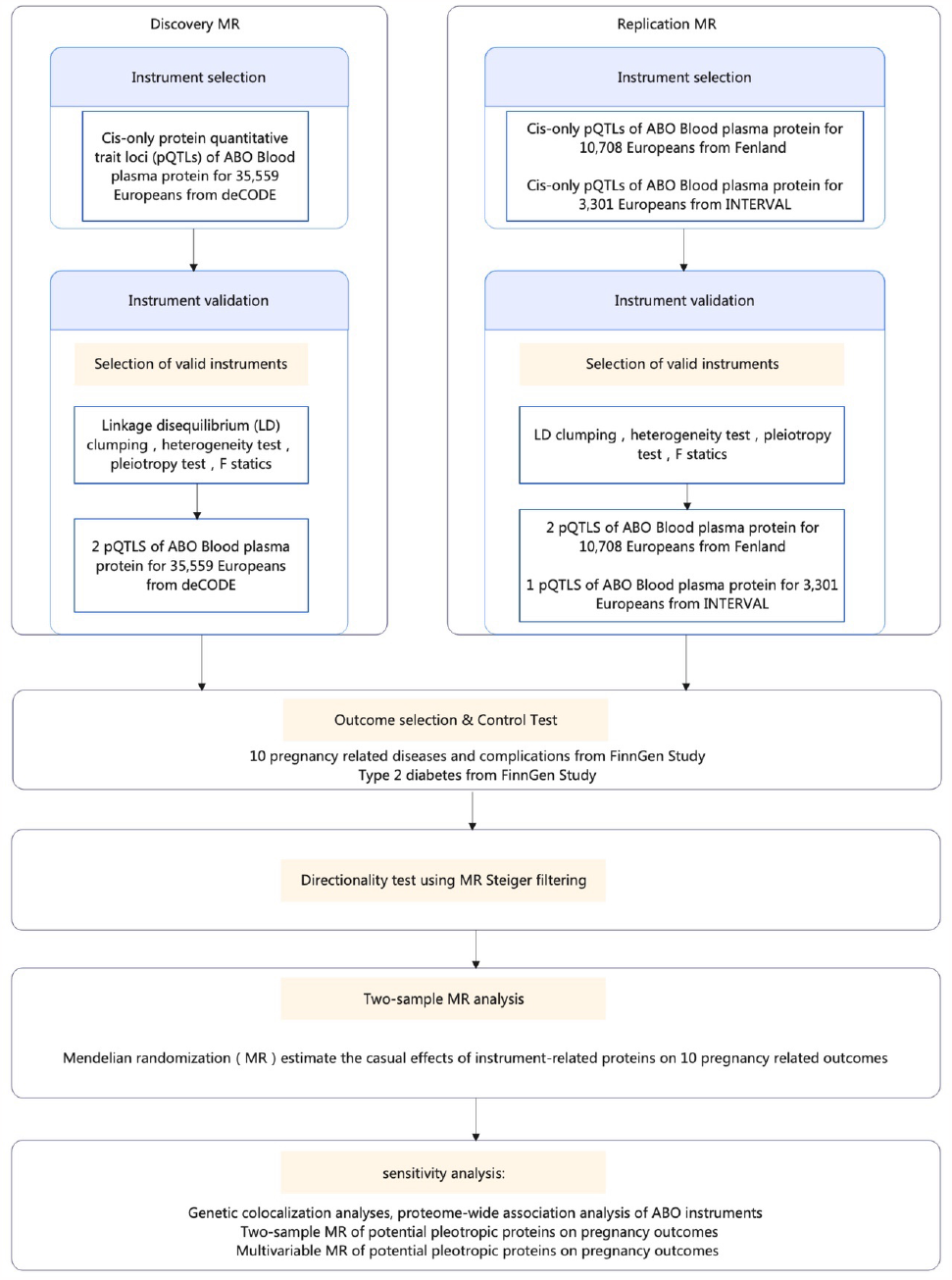
Study design of two-sample Mendelian randomization exploring effects of ABO protein with pregnancy outcomes Abbreviations: pQTL, protein quantitative trait loci; LD, linkage dise-quilibrium; T2D, type 2 diabetes.

**Figure 2.**
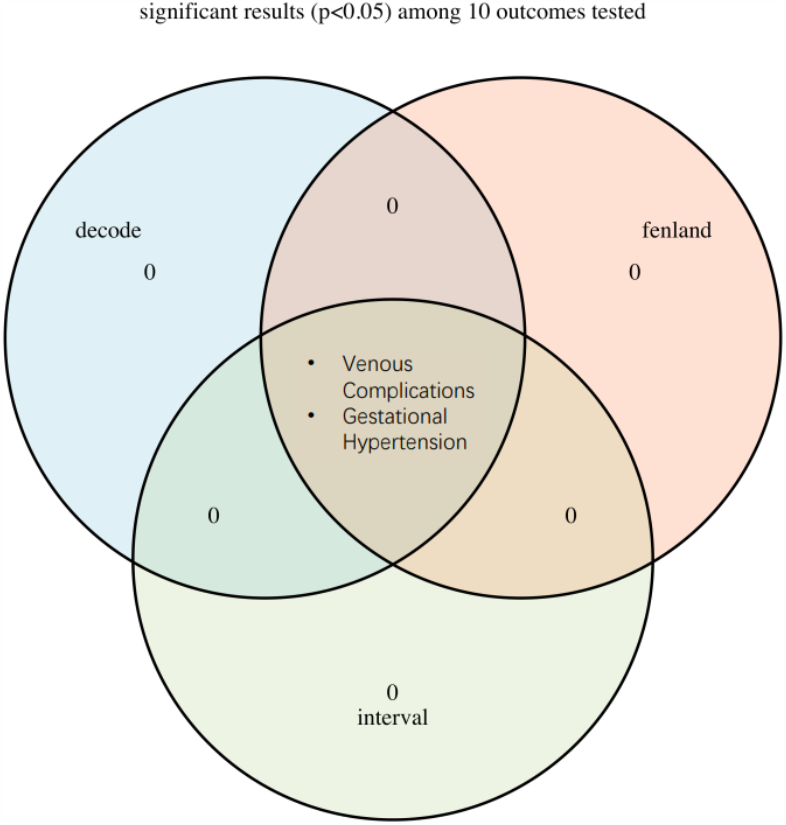
Venn diagram of Mendelian randomization estimates achieving statistical significance before multiple testing correction.

**Figure 3.**
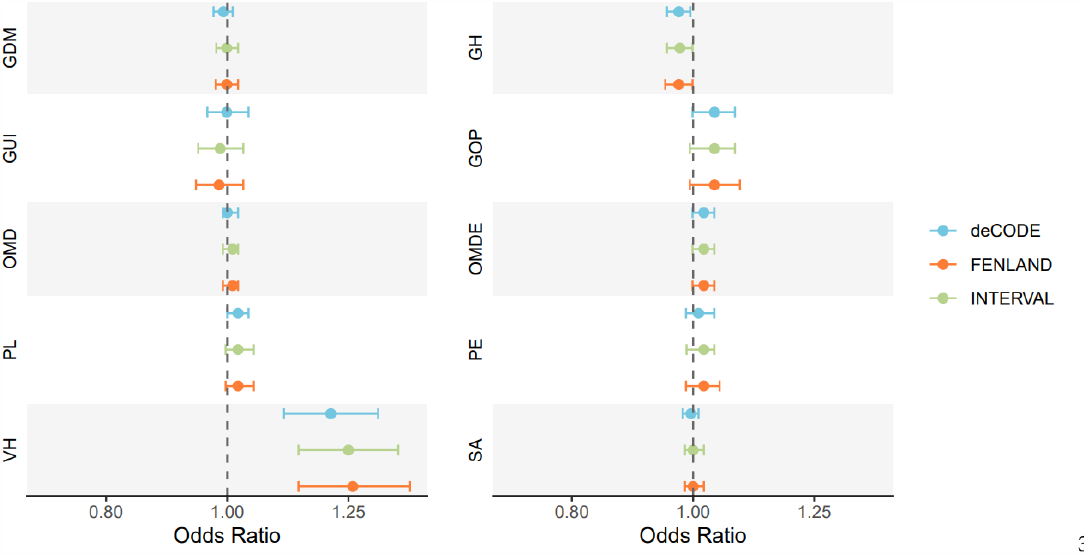
Two-sample Mendelian randomization estimates for associations of ABO protein with ten pregnancy outcomes Abbreviations: GDM, gestational diabetes mellitus; GH, gestational [pregnancy-induced] hypertension); GUI, infections of genitourinary tract in pregnancy; GOP, gestational [pregnancy-induced] oedema and proteinuria without hypertension; OMD, other maternal disorders predominantly related to pregnancy; OMDE, other maternal diseases classifiable else-where but complicating pregnancy, childbirth and the puerperium; PL, preterm labour and delivery; PE, pre-eclampsia; VH, venous complications and haemorrhoids in pregnancy; SA, spontaneous abortion.

**Figure 4.**
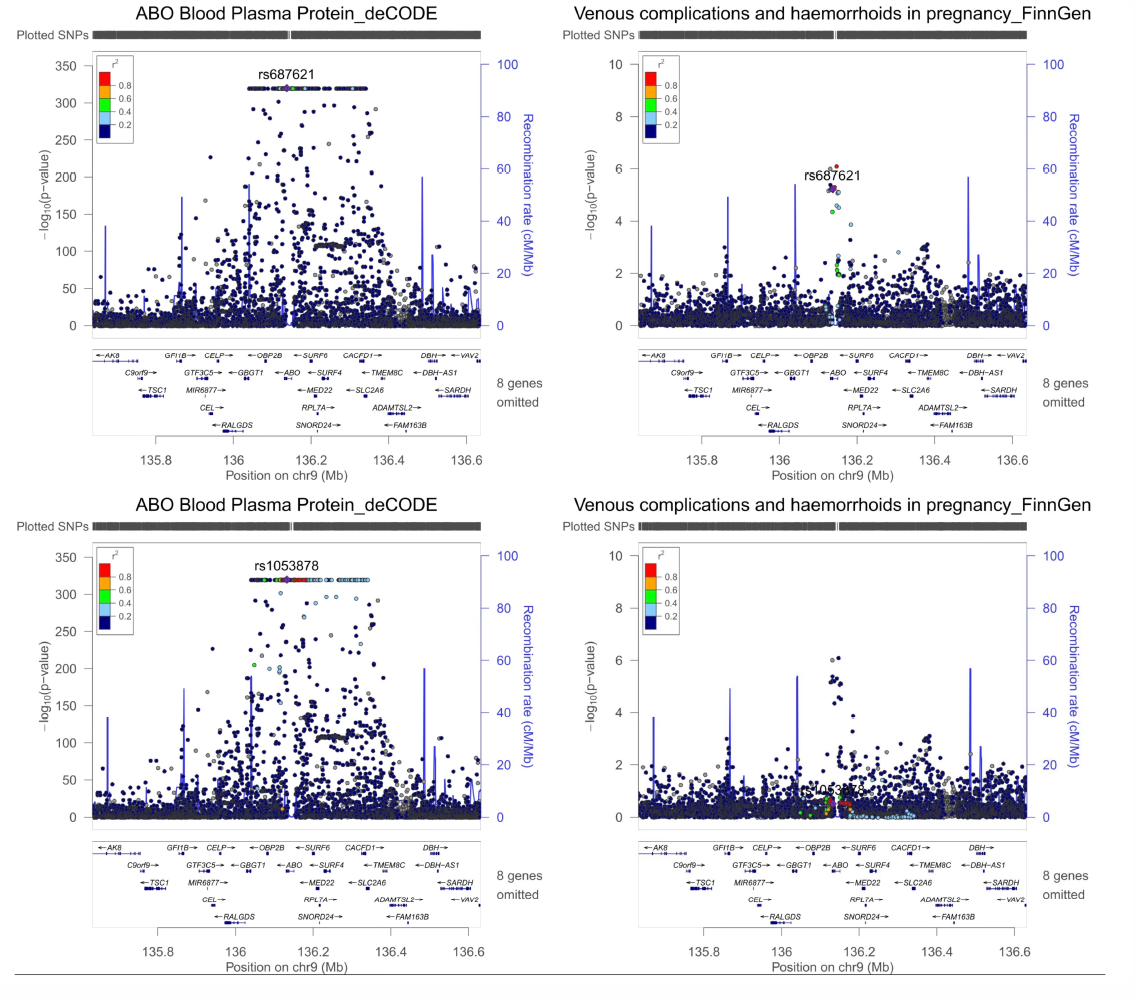
Regional plots of ABO pQTL – venous complications and haemorrhoids in pregnancy with Mendelian randomization evidence of potential causality.

### Effect of protein levels of ABO on pregnancy related outcomes using Fenland and INTERVAL data

In Table 1 and Figure 3, ABO pQTL was consistently associated with venous complications and haemorrhoids in pregnancy using data from Fenland and INTERVAL (OR = 1.263; 95% CI: 1.143–1.396; FDR-adjusted P-value = 5.001 x 10-5, OR = 1.248; 95% CI: 1.135– 1.372; FDR-adjusted P-value = 4.782 x 10-5, respectively). These estimates were also consistent with findings from the discovery analysis, suggesting the robustness of the causal relationship between ABO and venous complications. Consistent with the discovery MR, associations of ABO pQTL with other pregnancy related outcomes did not reach our FDR threshold here (Table 1 and Figure 3).

### Positive control analysis of protein levels of ABO on T2D

A higher protein level of ABO was associated with an increased risk of T2D using data from deCODE (OR = 1.038, 95%CI: 1.028–1.048, P = 9.421×10-15), Fenland (OR = 1.047, 95% CI: 1.036–1.059, P-value = 1.506×10-16) and INTERVAL (OR = 1.044, 95%CI: 1.034– 1.055, P-value = 2.091 x 10-16). Colocalization analysis also supports the causal effect of ABO pQTL on T2D (colocalization probability = 95.3%; Table1).

### Proteome-wide association study of ABO instruments and MR of potential pleiotropy proteins on pregnancy related outcomes

To estimate the influence of horizontal pleiotropy on the MR findings, we first looked up the genetic associations of ABO instruments on all available proteome GWAS listed in the IEU OpenGWAS database (prot-a,prot-b and prot-c; Table S8 and S9). We found that the two ABO instruments from deCODE (i.e., rs1053878 and rs950529388 (merged into rs687621)) were associated with 108 proteins in total, besides ABO protein. For these 108 proteins, we extracted their instruments and conducted two-sample MR of these proteins on our pregnancy related outcomes. We found that none of these proteins was associated with venous complications and haemorrhoids in pregnancy with Bonferroni-adjusted P-values < 0.01, which implies that unbalanced horizontal pleiotropy is less likely to be an issue for our MR results. We also identified that two proteins (toll-like receptor 4: lymphocyte antigen 96 complex and lactase-phlorizin hydrolase) were associated with GDM and GH, respectively (Bonferroni-adjusted P-values <0.01, Table S10). Using multivariable MR to adjust for these two proteins, we found little evidence for the association of ABO protein with GDM or GH (Table S11 and S12), which were consistent with our univariable MR results in Table 1.

## Discussion

In this study, we estimated the causal relationships of protein levels of ABO on ten pregnancy related outcomes using two-sample MR, and validated the findings in three independent datasets of ABO pQTL (deCODE as discovery, Fenland and INTERVAL as validation). Our study, for the first time, showed that one standard deviation higher of protein level of ABO was associated with an increased risk of pregnancy induced venous complications and haemorrhoids in pregnancy, while the other 55 proteins showed little evidence for an effect with this outcome. Sensitivity analyses including genetic colocalization and proteome-wide MR of potential pleiotropic instruments further strength the possibility of identifying true causal effects in this finding.

### Comparison with previous studies

In order to compare our top findings with previous literature, we comprehensively reviewed both conventional observational studies and MR studies estimating associations of ABO with any health outcomes. However, we did not identify any specific articles exploring an effect of circulating ABO protein on pregnancy related diseases or symptoms. Most existing studies focused on ABO blood group types, and found some associations with not only pregnancy outcomes (discussed below), but also outcomes in general population (e.g. cardiovascular diseases[26], knee osteoarthritis[27], stroke[28], thrombosis[29-31] and T2D[5]).

Our novel finding for the positive association of ABO protein expression in plasma with venous complications and haemorrhoids in pregnancy might not be directly compared to the previous studies investigating ABO blood group among the general population. Previous studies consistently showed that non-O blood group individuals had a significantly increased risk of venous thrombosis compared to O blood group individuals (i.e., non-ABO protein secretion), and the risk of thrombosis is likely to be inversely correlated with the expression of ABH antigens[29-31]. Additionally, consistent with our findings among pregnant women, ABO pQTL was found to be associated with venous thromboembolism in non-pregnant population[16]. Such evidence suggests that the ABO protein could be a significant new drug target to prevent venous complication during pregnancy by regulating the expression of ABH antigen.

Hypertensive disorders of pregnancy constitute one of the leading causes of maternal and perinatal mortality worldwide[32], and have been reported closely relating to ABO blood groups[33]. Our study provided some evidence to further support the casual relationship between ABO protein and gestational hypertension in the discovery analysis, although the findings did not pass FDR tests in the three datasets. It is worth mentioning that ABO plasma protein level and gestational hypertension across the three datasets showed similar effect estimates, suggesting that a higher level of ABO plasma protein might probably reduce the risk of gestational hypertension. Combing our finding for the unfavorable associations with venous complications, further studies need to confirm the protective effect on gestational hypertension, so as to inform antenatal care services to keep the ABO plasma protein within a proper range.

Previous observational studies showed conflicted results for the association of ABO level pre-eclampsia[34, 35], and our study provided little evidence for an association of ABO plasma protein level with pre-eclampsia. GDM is supposed to be closely related to T2D, and genome-wide association studies showed multiple lines of evidence pointed to the shared pathophysiology of GDM and T2D[36]. Recent studies also suggested that O blood group was independently associated with a higher risk of GDM[37], and AB blood group was considered as a protective factor[38]. Although we confirmed the causal relationship between ABO plasma protein level and T2D, there was little evidence supporting a causal relationship between ABO plasma protein level and GDM in our study. This could be due to insufficient power of the GDM GWAS, potential horizontal pleiotropy via other proteins, or suggests that the occurrence of T2D and GDM may be regulated by slightly different mechanisms.

### Strength and limitation

Key strengths of this study include that 1) to the best of our knowledge, it is the first study to use MR to explore associations of ABO protein with pregnancy outcomes; 2) comprehensive sensitivity analyses (e.g., positive control outcome, Steiger filtering, colocalization and proteome-wide MR) were applied to minimize potential bias due to violations of MR assumptions; 3) we investigated a range of adverse pregnancy outcomes in one paper.

However, limitations do exist. First, this study may be vulnerable to insufficient statistical power, as the numbers of cases ranged between 478 to 30971 for our outcomes. Although we identified statistically significant association with venous complications (the outcome with the smallest N cases), true causal effects on other outcomes (e.g., GDM) may be too small to be detect under the current sample sizes. Second, FinnGen defined venous symptoms and haemorrhoids in pregnancy (O22 in ICD-10) by combining multiple subtypes (O22.0 to O22.9 in ICD-10). Given we used summary-level data, we could not explore associations of ABO protein expression with those subtypes separately. Additionally, we noticed that both FinnGen and UK Biobank (field ID: 47270 and 47271) had very limited numbers of haemorrhoids cases based on their record linkage data of secondary care, even if this is expected to be a common disease in pregnancy[2]. This inconsistency could be because most patients did not have haemorrhoids severe enough to access to inpatient services. Thus, our results should be interpreted as the causal effect on severe venous conditions. Further studies based on self-report or primary care data are needed to investigated common diseases in pregnancy. Third, FinnGen included women with preexisting disorders (e.g., diabetes and hypertension) into the case groups, and did not exclude never pregnant women from the controls. Fourth, our genetic instruments were obtained from non-pregnant population, and we have to assume that the SNP-exposure associations are not varied during pregnancy. Fifth, our MR results may still be prone to horizontal pleiotropy due to paternal or fetal genotype[39]. We did not adjust for them because of limited publicly available GWAS data. Finally, our results are limited to women of European descent, and we cannot assume that they generalize to other populations.

## Conclusions

Our results suggest that plasma ABO protein level was causally associated with a higher risk of venous complications and hemorrhoids in pregnancy, which was unlikely to be influenced by horizontal pleiotropy. Our findings support prioritising ABO as a potential drug target for T2D and the pregnancy complications. However, we acknowledge the needs for further MR studies based on larger GWAS of pregnancy outcomes, large observational studies, and studies in women of non-European ancestries.

## Materials and Methods

### Genetic instruments selection of ABO protein levels

Figure 4 presented the study design. Cis-acting protein quantitative trait loci (pQTLs) from three proteome GWAS studies were used as candidate instruments for the discovery (deCODE n = 35,559) and validation (Fenland n = 10,708 and INTERVAL n = 3,301) MR analyses. The following criteria were applied for the instrument selection:

a. we only selected cis-acting pQTLs (defined as genetic variants ≤ 500 kb from the leading pQTL within the ABO region) since cis-acting pQTLs are more likely to have direct biological functions;
b. we selected pQTLs that were robustly associated with protein level of ABO (P-value threshold < 5 x 10-8);
c. we conducted linkage disequilibrium (LD) clumping for the instruments to identify pQTLs to proxy protein level of ABO in MR using generalized inverse variance weighted (gIVW) (threshold of LD r2 < 0.3 for exclusion of highly correlated pQTLs).

After selection, three groups of cis-acting pQTLs (two from deCODE [Table S1]; two from Fenland [Table S2]; one from INTERVAL [Table S3]) were used as instruments for the subsequent MR analyses (LD matrix among instruments were listed in [Table S4A, 4B and 4C]); Instruments from deCODE were selected for discovery analyses since it has the largest sample size among the three datasets, and the other two were used for validation analyses. The strength of genetic instruments was evaluated using F-statistic, with an F-statistic>10 suggesting a strong instrument[40].

### Genetic associations of pregnancy outcomes

A total of ten pregnancy related diseases and complications were selected as outcomes for this study, including venous complications and hemorrhoids in pregnancy, gestational hypertension, pre-eclampsia, infections of genitourinary tract in pregnancy, gestational oedema and proteinuria without hypertension, preterm labor and delivery, spontaneous abortion, gestational diabetes mellitus (GDM), and other outcomes related to childbirth(Table S5). Type 2 diabetes (T2D) was selected as a positive control outcome since its association with protein level of ABO has already been confirmed (Table S6)[41]. To avoid sample overlap with pQTL GWAS, we obtained GWAS summary statistics (including estimates of regression coefficient, the corresponding standard error and P value, effect allele, other allele, and effect allele frequency) for all the outcomes from FinnGen[19].

### Steiger filtering

With large sample sizes of the exposure and outcome GWAS data, we are likely to identify genetic variants robustly associated with both the exposure and outcomes. Therefore, we may observe reverse causality of genetic variants that influence the outcome first and then influence the exposure as a consequence. To minimise the possibility of reverse causality, we performed Steiger filtering[21] to select valid genetic instruments that showed a right direction of exposure-outcome effect. Steiger filtering assumes that a valid instrument should explain more variance of the exposure than that of the outcome, and identifies those instruments that explained more variance of the outcome than that of the exposure for removal from MR analyses.

### MR analysis

We applied two-sample MR to estimate the effect of protein level of ABO on pregnancy outcomes (βXY). Wald ratio estimate[42], i.e., the ratio of the genetic association of the outcome (βGY) against the genetic association of the exposure (βGX), was used to estimate the effect based on each instrument. The gIVW with multiplicative random effects[43], was applied to combine the Wald ratio estimates for multiple variants for the same exposure. We conducted the discovery MR by using genetic instruments from deCODE, and re-conducted gIVW by using those from INTERVAL and Fenland for validation. Since these instruments are obtained from three independent European-based cohorts, the reliability of our MR findings will increase if consistent results are shown across those datasets. To control for multiple testing, we used Benjamini-Hochberg method to calculate the false discovery rate (FDR) (threshold FDR-adjusted P-value<0.05) for robust results.

### Colocalization analyses

We conducted genetic colocalization analyses to detect whether the identified MR signals shared causal variants between the protein expression of ABO and pregnancy outcomes, or were just due to confounding by LD. Under the assumption of a single causal variant within each genomic region, the Bayesian statistical framework quantifies the posterior probability of association (PPA) for each of the four possible hypotheses[23] in validation analysis (H1/H2: the genetic variants [i.e., pQTLs] were only associated with the protein level of ABO or associated with a pregnancy outcome in the tested genomic region; H3: there were genetic variants associated with both protein expression of ABO and a pregnancy outcome in the tested genomic region but the two traits did not shared the same casual variant; H4: there were genetic variants associated with both protein expression of ABO and a pregnancy outcome, and the two traits shared the same casual variant in the tested genomic region). The effect estimates and allele information for all variants within 500KB window upstream or downstream of the leading pQTL for each trait were extracted for the colocalization analysis. We assigned prior probabilities that a variant is associated with trait 1 (p1 = 1 x 10-4), trait 2 (p2 = 1 x 10-4), and both traits (p12 = 1 x 10-5) as recommended. The PP H4/(PP H3+ PP H4) ≥ 70% was considered as the threshold for colocalization tests. We also conducted an LD check analysis between ABO pQTL and the outcome that had showed evidence for colocalizaton using the above Bayesian method, to further validate the results. The LD check window was set up as ±500kb, and LD r2 ≥ 0.8 was considered as evidence for colocalization[20].

### Proteome-wide association analysis of ABO instruments and MR of potential pleiotropic proteins on pregnancy outcomes

It is known that the ABO pQTLs were pleiotropic, being associated with a set of proteins[16]. To evaluate the influence of their horizontal pleiotropy, we performed a two-step proteome-wide MR approach using the instruments from deCODE. First, we searched for the genetic effects of each ABO instrument across all publicly available proteome-wide GWAS on IEU OpenGWAS[17, 18] to identify its effect on other proteins. These were potential pleiotropic proteins that may link ABO instruments with outcomes not via ABO protein expression. Then we selected those proteins that were associated with our instruments as exposures, and used MR method to check whether there were casual relationships between those potential pleiotropic proteins and our pregnancy outcomes. All pQTLs associated with these potential pleiotropic proteins were used as instruments for this analysis. To control for multiple testing, we used the conservative Bonferroni-adjusted P-value (i.e., P ≤ 0.05) to define robust MR estimates. Where a pleiotropic protein identified for an outcome, we further conducted a multivariable MR to obtain the direct effect of ABO protein on the outcome adjusting for the pleiotropic protein[44].

## Data Availability

All data produced in the present study are available upon reasonable request to the authors.

## Author Contributions

J. Zheng, Y. Bi and H. Zhao contributed to the study conception and design. Material preparation and data collection were performed by R. Gu, M. Wang and J. Zheng; Analyses were performed by H. Zheng, Y. Sun and M. Wang; the manuscript was written by Y. Sun, H. Zheng, R. Gu, M. Wang, X. Wu and Q. Yang, with critical comments from Y. Bi, J. Zheng, H. Zhao, and Q. Yang. All authors commented on previous versions of the manuscript. All authors have read and agreed to the published version of the manuscript.

## Data Availability Statement

This study only used publicly available GWAS summary statistics from deCODE, Fenland, INTERVAL, FinnGen (https://www.finngen.fi/en) and IEU OpenGWAS (https://gwas.mrcieu.ac.uk/).

## Acknowledgments

The authors would like to thank deCODE, Fenland, INTERVAL and FinnGen investigators for sharing their summary-level data.

## References

1. Goldenberg, R.L., et al., Epidemiology and causes of preterm birth. Lancet, 2008. 371(9606): p. 75–84.

2. Alsheef, M.A., et al., Pregnancy and Venous Thromboembolism: Risk Factors, Trends, Management, and Mortality. Biomed Res Int, 2020. 2020: p. 4071892.

3. Vazquez, J.C., Constipation, haemorrhoids, and heartburn in pregnancy. BMJ Clin Evid, 2010. 2010.

4. Kintiraki, E., et al., Pregnancy-Induced hypertension. Hormones (Athens), 2015. 14(2): p. 211–23.

5. Abegaz, S.B., Human ABO Blood Groups and Their Associations with Different Diseases. Biomed Res Int, 2021. 2021: p. 6629060.

6. Hoglund, J., et al., Characterization of the human ABO genotypes and their association to common inflammatory and cardiovascular diseases in the UK Biobank. Am J Hematol, 2021. 96(11): p. 1350–1362.

7. Yu, J.Y., et al., Relationship between maternal ABO blood groups and pregnancy outcomes: a retrospective cohort study in Dongguan, China. Journal of Obstetrics and Gynaecology, 2023. 43(2).

8. Rizzo, A.N. and E.P. Schmidt, ABO blood type: a window into the genetics of acute respiratory distress syndrome susceptibility. J Clin Invest, 2021. 131(1).

9. Franchini, M., C. Mengoli, and G. Lippi, Relationship between ABO blood group and pregnancy complications: a systematic literature analysis. Blood Transfus, 2016. 14(5): p. 441–8.

10. Committee on, E., ACOG Committee Opinion No. 646: Ethical Considerations for Including Women as Research Participants. Obstet Gynecol, 2015. 126(5): p. e100–7.

11. Smith, G.D. and S. Ebrahim, ‘Mendelian randomization’: can genetic epidemiology contribute to understanding environmental determinants of disease? Int J Epidemiol, 2003. 32(1): p. 1–22.

12. Luo, S., et al., Identifying factors contributing to increased susceptibility to COVID-19 risk: a systematic review of Mendelian randomization studies. Int J Epidemiol, 2022. 51(4): p. 1088–1105.

13. Chong, M., et al., Novel Drug Targets for Ischemic Stroke Identified Through Mendelian Randomization Analysis of the Blood Proteome. Circulation, 2019. 140(10): p. 819–830.

14. Halldorsson, B.V., et al., The sequences of 150,119 genomes in the UK Biobank. Nature, 2022. 607(7920): p. 732–740.

15. Pietzner, M., et al., Mapping the proteo-genomic convergence of human diseases. Science, 2021. 374(6569): p. eabj1541.

16. Sun, B.B., et al., Genomic atlas of the human plasma proteome. Nature, 2018. 558(7708): p. 73–79.

17. Ben Elsworth, M.L., Tessa Alexander, Yi Liu, Peter Matthews, Jon Hallett, Phil Bates, Tom Palmer, Valeriia Haberland, George Davey Smith, Jie Zheng, Philip Haycock, Tom R Gaunt, Gibran Hemani., The MRC IEU OpenGWAS data infrastructure. bioRxiv

18. Hemani, G., et al., The MR-Base platform supports systematic causal inference across the human phenome. Elife, 2018. 7.

19. Kurki, M.I., et al., FinnGen provides genetic insights from a well-phenotyped isolated population. Nature, 2023. 613(7944): p. 508-+.

20. Zheng, J., et al., Phenome-wide Mendelian randomization mapping the influence of the plasma proteome on complex diseases. Nat Genet, 2020. 52(10): p. 1122–1131.

21. Hemani, G., K. Tilling, and G. Davey Smith, Orienting the causal relationship between imprecisely measured traits using GWAS summary data. PLoS Genet, 2017. 13(11): p. e1007081.

22. Richardson, T.G., et al., Mendelian Randomization Analysis Identifies CpG Sites as Putative Mediators for Genetic Influences on Cardiovascular Disease Risk. Am J Hum Genet, 2017. 101(4): p. 590–602.

23. Giambartolomei, C., et al., Bayesian test for colocalisation between pairs of genetic association studies using summary statistics. PLoS Genet, 2014. 10(5): p. e1004383.

24. Barbeira, A.N., et al., Exploring the phenotypic consequences of tissue specific gene expression variation inferred from GWAS summary statistics. Nat Commun, 2018. 9(1): p. 1825.

25. Boughton, A.P., et al., LocusZoom.js: interactive and embeddable visualization of genetic association study results. Bioinformatics, 2021. 37(18): p. 3017–3018.

26. Chen, Y., et al., Analysis of circulating cholesterol levels as a mediator of an association between ABO blood group and coronary heart disease. Circ Cardiovasc Genet, 2014. 7(1): p. 43–8.

27. Li, C., et al., Association between the ABO blood group and primary knee osteoarthritis: A case-control study. J Orthop Translat, 2020. 21: p. 129–135.

28. Lotz, R.C., et al., ABO blood group system and occurrence of ischemic stroke. Arq Neuropsiquiatr, 2021. 79(12): p. 1070–1075.

29. Clark, P. and O. Wu, ABO blood groups and thrombosis: a causal association, but is there value in screening? Future Cardiol, 2011. 7(2): p. 191–201.

30. Franchini, M. and P.M. Mannucci, ABO blood group and thrombotic vascular disease. Thromb Haemost, 2014. 112(6): p. 1103–9.

31. Franchini, M. and G. Lippi, Relative Risks of Thrombosis and Bleeding in Different ABO Blood Groups. Semin Thromb Hemost, 2016. 42(2): p. 112–7.

32. Gestational Hypertension and Preeclampsia: ACOG Practice Bulletin Summary, Number 222. Obstet Gynecol, 2020. 135(6): p. 1492–1495.

33. Lee, B.K., et al., ABO and RhD blood groups and gestational hypertensive disorders: a population-based cohort study. BJOG, 2012. 119(10): p. 1232–7.

34. Clark, P. and O. Wu, ABO(H) blood groups and pre-eclampsia. A systematic review and meta-analysis. Thromb Haemost, 2008. 100(3): p. 469–74.

35. Hentschke, M.R., et al., Is there any relationship between ABO/Rh blood group and patients with pre-eclampsia? Pregnancy Hypertens, 2014. 4(2): p. 170–3.

36. Pervjakova, N., et al., Multi-ancestry genome-wide association study of gestational diabetes mellitus highlights genetic links with type 2 diabetes. Hum Mol Genet, 2022. 31(19): p. 3377–3391.

37. Sapanont, K., P. Sunsaneevithayakul, and D. Boriboonhirunsarn, Relationship between ABO blood group and gestational diabetes mellitus. Journal of Maternal-Fetal & Neonatal Medicine, 2021. 34(8): p. 1255–1259.

38. Rom, E., et al., The association between ABO blood groups and gestational diabetes mellitus: a retrospective population-based cohort study. Journal of Maternal-Fetal & Neonatal Medicine, 2022. 35(25): p. 7065–7069.

39. Lawlor, D., et al., Using Mendelian randomization to determine causal effects of maternal pregnancy (intrauterine) exposures on offspring outcomes: Sources of bias and methods for assessing them. Wellcome Open Res, 2017. 2: p. 11.

40. Pierce, B.L., H. Ahsan, and T.J. Vanderweele, Power and instrument strength requirements for Mendelian randomization studies using multiple genetic variants. Int J Epidemiol, 2011. 40(3): p. 740–52.

41. Getawa, S., et al., Relationships of ABO and Rhesus blood groups with type 2 diabetes mellitus: a systematic review and meta-analysis. J Int Med Res, 2022. 50(10): p. 3000605221129547.

42. Burgess, S., F. Dudbridge, and S.G. Thompson, Combining information on multiple instrumental variables in Mendelian randomization: comparison of allele score and summarized data methods. Stat Med, 2016. 35(11): p. 1880–906.

43. Bowden, J., et al., Consistent Estimation in Mendelian Randomization with Some Invalid Instruments Using a Weighted Median Estimator. Genet Epidemiol, 2016. 40(4): p. 304–14.

44. Sanderson, E., et al., An examination of multivariable Mendelian randomization in the single-sample and two-sample summary data settings. Int J Epidemiol, 2019. 48(3): p. 713–727.

